# Evaluation of the Panbio™ COVID-19 Antigen Rapid Diagnostic Test in subjects infected with Omicron using different specimens

**DOI:** 10.1101/2022.03.19.22272637

**Authors:** Rafael Mello Galliez, Larissa Bomfim, Diana Mariani, Isabela de Carvalho Leitão, Anna Carla Pinto Castiñeiras, Cassia Cristina Alves Gonçalves, Bianca Ortiz da Silva, Pedro Henrique Cardoso, Monica Barcelos Arruda, Patricia Alvarez, Victor Akira Ota, Débora Gomes Marins Rodrigues, Luciana Jesus da Costa, Orlando da Costa Ferreira, Terezinha Marta Pereira Pinto Castiñeiras, Debora Souza Faffe, Amilcar Tanuri

**Affiliations:** Núcleo de Enfrentamento e Estudos de Doenças Infecciosas Emergentes e Reemergentes, Universidade Federal do Rio de Janeiro, Rio de Janeiro, Brasil; Laboratório de Virologia Molecular, Departamento de Genética, Instituto de Biologia, Universidade Federal do Rio de Janeiro, Rio de Janeiro, Brasil; Instituto de Biofísica Carlos Chagas Filho, Universidade Federal do Rio de Janeiro, Rio de Janeiro, Brasil; Instituto de Microbiologia Paulo de Góes, Universidade Federal do Rio de Janeiro, Rio de Janeiro, Brasil; Departamento de Doenças Infecciosas e Parasitárias, Faculdade de Medicina, Universidade Federal do Rio de Janeiro, Rio de Janeiro, Brasil; Hospital Universitário Clementino Fraga Filho, Centro de Ciências da Saúde, Universidade Federal do Rio de Janeiro, Rio de Janeiro, Brasil; Decania, Centro de Ciências da Saúde, Universidade Federal do Rio de Janeiro, Rio de Janeiro, Brasil; Instituto de Tecnologia de Imunobiológicos Bio-manguinhos, Fundação Oswaldo Cruz/Fiocruz, Rio de Janeiro, Brasil

## Abstract

Community testing is a crucial tool for the early identification of SARS-CoV-2 infection and transmission control. The emergence of the highly mutated Omicron variant (B.1.1.259) raised concerns about its primary site of replication, impacting sample collection, and its detectability by rapid antigens tests. We tested the Antigen Rapid Diagnostic Test (Ag-RDT) performance using nasal, oral, and saliva specimens for COVID-19 diagnosis in 192 symptomatic individuals, using RT-qPCR from nasopharyngeal samples as control. Variant of Concern (VOC) investigation was determined by the 4Plex SARS-CoV-2 screening kit. SARS-CoV-2 positivity rate was 66.2%, with 99% of the positive samples showing an amplification profile consistent with that of the Omicron variant. Nasal Ag-RDT showed higher sensitivity (89%) than oral (12.6%) and saliva (22.1%) Ag-RDTs. Our data showed the good performance of the Ag-RDT in a pandemic scenario dominated by the Omicron VOC. Furthermore, our data also demonstrated that nasal specimens perform better than oral and saliva ones for Omicron Ag-RDT detection.

## 1. Introduction

Since the beginning of the COVID-19 pandemic, the World Health Organization has advised widespread testing to identify infected individuals and control the onward transmission of the virus [1,2]. SARS-CoV-2 can spread quickly in the global population and new variants can emerge due to different selective pressures [3]. In fact, the prolonged circulation of the virus has resulted in the emergence of multiple variants in different countries during the COVID-19 pandemic. Some variants are of great interest to public health due to the critical mutations they carry in the Spike (S) protein. These mutations, named Variant of Concern (VOC), can change the binding of neutralizing antibodies as well as the affinity of S to the angiotensin-converting enzyme 2 (ACE2) receptor [4,5]. Since June 2021, we have faced several global waves of VOCs, such as Alpha, Beta, Gamma, Delta, and more recently Omicron [5,6].

The latest Omicron variant (B.1.1.259) replaced the Delta variant very quickly and recently dominated the pandemic worldwide [7]. Omicron is a highly mutated strain, including 50 mutations in its genome and at least 32 in the Spike protein. The following mutations are present in Omicron Spike protein (A67V, Δ69−70, T95I, G142D, Δ143-145, Δ211, L212I, insertion 214-EPE, G339D, S371L, S373P, S375F, K417N, N440K, G446S, S477N, T478K, E484A, Q493R, G496S, Q498R, N501Y, Y505H, T547K, D614G, H655Y, N679K, P681H, N764K, D796Y, N856K, Q954H, N969K and L981F). All these variations could impact Omicron’s ability to escape from monoclonal antibodies and from neutralizing antibodies elicited by COVID-19 vaccines [7,8]. Indeed, studies have reported approximately 25-fold to 40-fold reductions in serum neutralizing activity compared to historical D614G-containing strains from individuals immunized with the Pfizer BNT162b2 and AstraZeneca AZD1222 vaccines [9-13].

RT-qPCR is considered the gold-standard test in COVID-19 diagnosis to detect the viral genetic material in different body fluids [14]. SARS-CoV-2 starts its replication in the upper respiratory compartment making the nasopharynx the most informative site for swab collection since it is rich in viral RNA and antigens [15]. However, nasal swabs have also shown good sensitivity [16]. Other locations and fluids can also be used in COVID-19 molecular tests such as oral or gingival swabs and even spit saliva.

Whereas RT-qPCR plays an essential role in the detection of infected individuals, the Antigen Rapid Diagnostic Tests (Ag-RDT) arose as a tool to detect SARS-CoV-2 viral proteins in a faster and less expensive way [1]. Most Ag-RDTs use a monoclonal antibody directed against the Nucleocapsid (N) protein. Mutations in VOCs are frequently present at the S protein. However, they can also occur at non-structural and other structural proteins such as N or ORFs 3, 6, 7, and 8 [4,5]. The major concern is that VOC-related mutations can disturb the binding of the N capture monoclonal antibodies and decrease the sensitivity of the test. Omicron has a unique mutation in N protein (P13L) and a deletion of two amino acids (ER) in positions 32-33 [6]. The impact of these N protein mutations in the binding of monoclonal antibodies responsible for the capture and development of the Ag-RDT is unknown. The other way VOCs can impact the Ag-RDT results is by changing the initial site of viral replication. Usually, swab specimens are collected from the nasopharynx or nasal cavity. If VOC viral replication initiates in the oral cavity instead, before spreading to the nasopharynx and nasal cavity, the usually collected specimens could fail to detect the initial phase of infection. This fact was reported by Marais et al 2021 during the beginning of Omicron spread in South Africa [17]. In the present study, we explored the diagnostic performance of nasal and oral swabs as well as saliva in symptomatic patients infected with the Omicron variant.

## 2. Materials and methods

### 2.1. Study design

Our main aim was to evaluate the performance of Ag-RDT on different specimens (nasal, oral, and saliva) compared with RT-qPCR from the nasopharyngeal swab as the gold-standard. Our secondary aim was to analyze the persistence/disappearance of Ag-RDT positivity in the different specimens until the 7th day after a positive diagnosis by RT-qPCR.

### 2.2. Ethical statement

The present study was approved by the local ethics review committee from Clementino Fraga Filho University Hospital (CAAE: 30161620.0.0000.5257) and by the national ethical review board (CAAE: 30127020.0.0000.0068). All enrolled participants signed written informed consent.

### 2.3. Study population

Subjects were over 18 years old and presented with mild COVID-19 suggestive symptoms -such as fever, cough, runny nose, sore throat, anosmia, ageusia, headache, diarrhea, and myalgia-for up to 7 days.

### 2.4 Sample collection

A trained healthcare professional collected four specimens from each participant. Samples included a nasal bilateral middle-turbinate swab, an oral swab, a saliva sample, and a nasopharyngeal swab. Nasal samples were collected according to the Panbio™ COVID-19 Ag Test Device instructions for use. Participants abstained from ingestion of food, drink, or chewing tobacco and gum for 30 minutes before oral sample collection. Participants coughed 3-5 times, covering their mouth with disposable tissue paper, prior to the operator swabbing the inside of both cheeks, above and below the tongue, on gums, and hard palate. Furthermore, participants spat in a sterile recipient to collect at least 1 mL of saliva, then 300 µL were transferred with a pipette to the Ag-RDT elution tube. After 5-10 minutes of rest, the operator collected a nasopharyngeal swab and immediately placed it in viral transport media (VTM).

### 2.5 Ag-RDT procedure

Nasal, oral, and saliva specimens were tested immediately following collection using the Panbio™ COVID-19 Ag Test Device (Abbott) according to the manufacturer’s instructions. Tests were run and read by trained technicians on-site in the testing center, within 15 minutes. The leftover samples of the Panbio™ elution tubes (∼120 µL) were stored at 4 °C and shipped to the laboratory for RT-qPCR within 4 hours.

### 2.6 Quantification of viral loads via RT-qPCR

Viral RNA from the nasopharyngeal swab, as well as nasal, oral, and saliva leftover samples were extracted in a KingFisher Flex System® (Thermofisher, USA), using the MagMax Viral/Pathogen Kit (Thermofisher, USA). SARS-CoV-2 (2019-nCoV) multiplex CDC qPCR Probe Assay (Integrated DNA Technologies, USA) targeting the SARS-CoV-2 N1 and N2 genes and the human ribonuclease P (RNaseP) gene (endogenous control) detected viral RNA. A 7500 Thermal Cycler (Applied Biosystems, USA) was used for all reactions. RT-qPCR results were interpreted as positive if both targets (N1 and N2) were amplified with cycle threshold (Ct) ≤ 37.

VOC investigations were made on SARS-CoV-2 RT-qPCR positive samples using a 4Plex SARS-CoV-2 for VOC screening kit (Bio-Manguinhos, Brazil). TaqMan probes for the SARS-CoV-2 virus were used for detection by amplifying a target region in the N gene and screening samples with suggestive profiles for the different VOCs. VOC profiles were given by combining results obtained of the deletions (Del) S106, G107, and F108 in the ORF1a gene (NSP6) and Del. H69 and V70 in the Spike gene from the samples tested. Samples were considered positive when Ct values for SC2-N, Wt Del NSP6, and Wt Del 69, 70 were lower than 40.

### 2.7 Statistical analysis

The data from the study was acquired using the KoBoCollect online/offline web-based form (Available at: https://www.kobotoolbox.org). The dataset was extracted on XLSForms and merged with the corresponding laboratory data. Ag-RDT sensitivity and specificity from different specimens were determined in relation to RT-qPCR. Sensitivity, specificity, predictive positive and negative values, as well as 95% confidence intervals (CI) were determined using Fisher’s exact test. Differences among groups were assessed by Kruskal-Wallis non-parametric test. GraphPad Prism version 9.2.0 (GraphPad Software, San Diego, California USA), and JASP Version 0.16 (JASP Team, 2021, Computer software) were used. A p-value < 0.05 was considered significant.

## 3. Results

From January 14, 2022, to February 7, 2022, a total of 192 individuals were tested for SARS-CoV-2 by RT-qPCR and Panbio™ Ag-RDT tests simultaneously at the Center for COVID-19 diagnosis of the Federal University of Rio de Janeiro. Table 1 describes the general characteristics of the study cohort. Females represented the majority of patients (65.6%), with a mean age of 39 years. Most individuals were tested within three days of symptoms onset (66.1%). Nearly all patients sampled were fully immunized against COVID-19 (97.9%), 67.2% with a 3rd vaccine shot.

**Table 1.** Cohort general characteristics

| **Characteristic** | **Total (%)** |
| --- | --- |
| **Number of patients** | 192 |
| **Age [average (min-max)]** | 39 (21-74) years old |
| **Gender** |  |
| Female | 126 (65.6) |
| Male | 66 (34.4) |
| **Days since symptom onset (DSSO) [mode (min-max)]** | 3 (0-7) |
| DSSO $\leq 3$ | 127 (66.1) |
| DSSO 4-7 | 65 (33.9) |
| **Vaccination** |  |
| 1 dose | 4 (2.1) |
| 2 doses | 59 (30.7) |
| $\geq 3$ doses | 129 (67.2) |
| **Follow-up** | 32 (16.7) |
| DSSO [mode (min-max)] | 10 (2-12) |
| **Positivity rate** |  |
| RT-qPCR | 127 (66.2) |
| Ag_Nasal | 113 (58.9) |
| Ag_Oral | 16 (8.6) |
| Ag_Saliva | 28 (14.6) |

The SARS-CoV-2 positivity rate in our cohort was higher using RT-qPCR from nasopharyngeal swabs (66.2%) than Ag-RDTs. Nasal specimens yielded the highest positivity rate among the different Ag-RDTs (58.9%) compared with oral (8.6%) and saliva (14.6%) (Table 1). The performance of Ag-RDT was higher when using nasal specimens than oral or saliva specimens, yielding 89% sensitivity compared with 12.6% and 22.1%, respectively. The specificity of Ag-RDT was 100% regardless of the specimen tested (Table 2).

**Table 2.** Sensitivity and specificity of Ag-RDT in relation to SARS-CoV-2 RT-qPCR result

|  | RT-qPCR |  |  | Sensitivity (95% CI) | Specificity (95% CI) | Positive predictive value (95% CI) | Negative predictive value (95% CI) |
| --- | --- | --- | --- | --- | --- | --- | --- |
|  | Positive | Negative | Total |  |  |  |  |
| Ag-RDT |  |  |  |  |  |  |  |
| Nasal |  |  |  |  |  |  |  |
| Positive | 113 | 0 | 113 | 89.0% (82.4-93.3) | 100.0% (94.4-100.0) | 100.0% (96.7-100.0) | 82.3% (72.4-89.1) |
| Negative | 14 | 65 | 79 |  |  |  |  |
| Total | 127 | 65 | 192 |  |  |  |  |
| Oral |  |  |  |  |  |  |  |
| Positive | 16 | 0 | 16 | 12.6% (7.9-19.5) | 100.0% (94.4-100.0) | 100.0% (80.6-100.0) | 36.9% (30.2-44.3) |
| Negative | 111 | 65 | 176 |  |  |  |  |
| Total | 127 | 65 | 192 |  |  |  |  |
| Saliva |  |  |  |  |  |  |  |
| Positive | 28 | 0 | 28 | 22.1% (15.7-30.0) | 100.0% (94.4-100.0) | 100.0% (87.9-100.0) | 39.6% (32.5-47.3) |
| Negative | 99 | 65 | 164 |  |  |  |  |
| Total | 127 | 65 | 192 |  |  |  |  |

When we analyzed the concordance among RT-qPCR and Ag-RDT results from different specimens, nine possible results were obtained (Figure 1). Most patients positive for SARS-CoV-2-RT-qPCR (n = 127) were also positive for nasal Ag-RDT (n = 113), while only 16 and 28 were detected by Ag-RDT from oral and saliva samples, respectively. The nasal Ag-RDT result showed the best concordance with RT-qPCR (n = 82), while Ag-RDT from oral and saliva samples alone failed to detect patients with positive SARS-CoV-2 RT-qPCR results (Figure 1).

**Figure 1.**
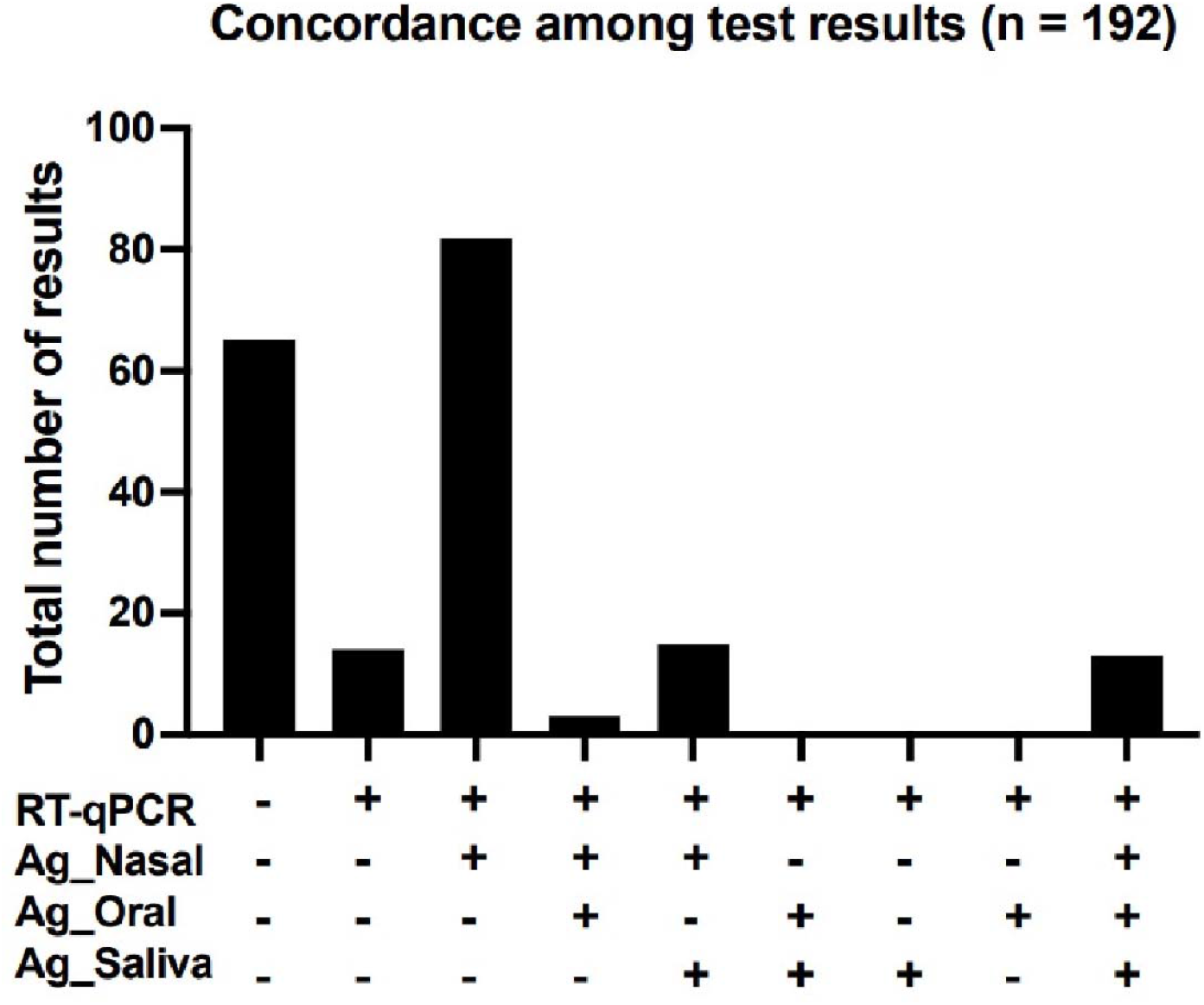
Concordance among SARS-CoV-2 RT-qPCR and Ag-RDT results from nasal (Ag_Nasal), oral (Ag_Oral), and saliva (Ag_Saliva) specimens from 192 mildly symptomatic patients analyzed up to 7 days since symptom onset.

Median cycle threshold (Ct) values of N1 target amplification by RT-qPCR from nasopharyngeal samples and Ag-RDT leftovers from nasal, oral, and saliva specimens were significantly different (Figure 2). The median Ct values were higher in nasopharyngeal samples and nasal Ag-RDT leftovers than those obtained from oral or saliva Ag-RDTs.

**Figure 2.**
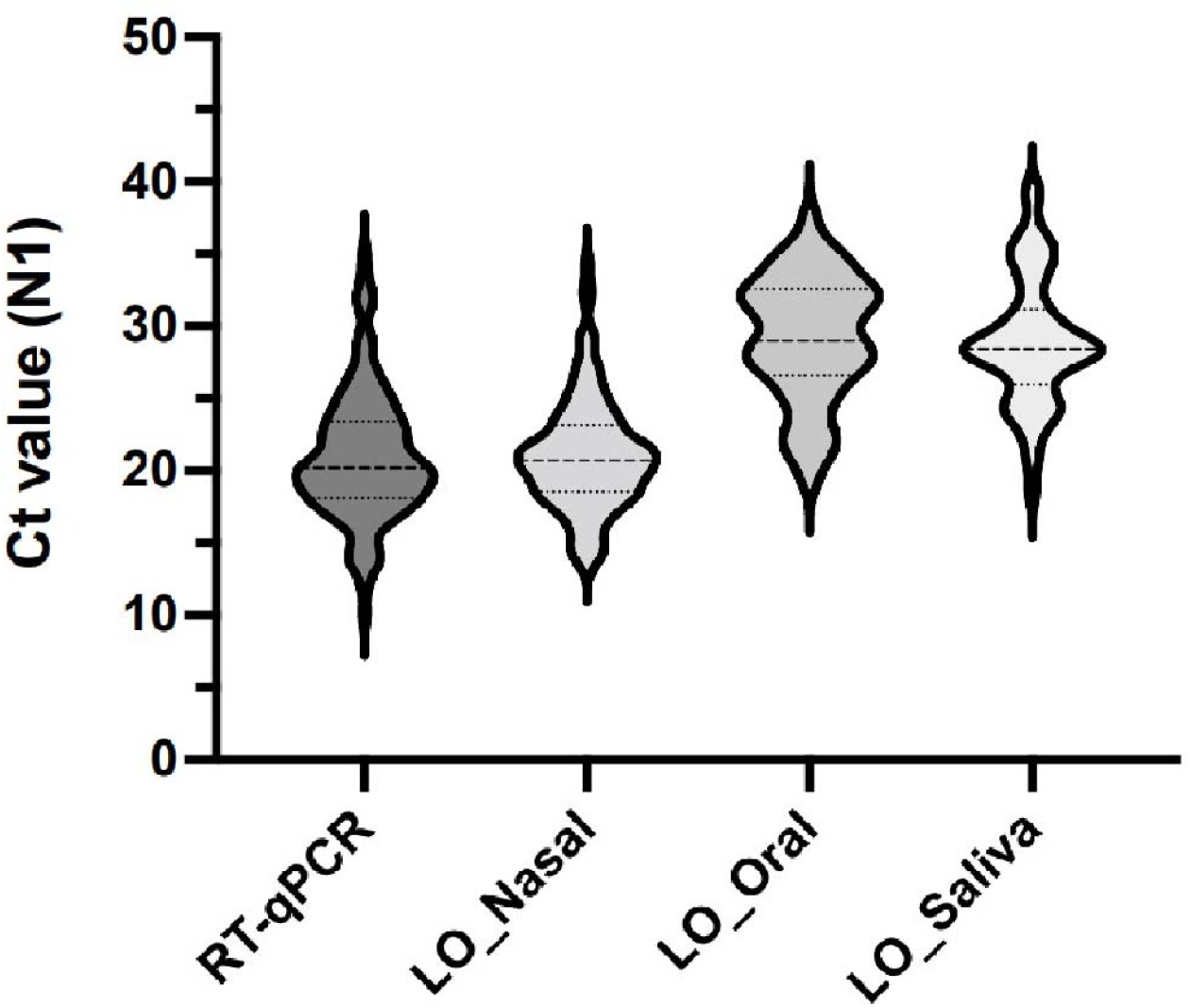
Violin plots showing the median, variability, and probability density of cycle threshold (Ct) values of N1 target amplification obtained by RT-qPCR from nasopharyngeal samples (RT-qPCR) and Ag-RDT leftovers (LO) from nasal, oral, and saliva specimens in 192 mildly symptomatic patients tested up to 7 days since symptom onset. Medians were significantly different among groups (Kruskal-Wallis non-parametric test, *p* = 0.0009).

Ninety-seven specimens with positive SARS-CoV-2 RT-qPCR results and Ct value below 30 were segregated to run the VOC RT-qPCR. Ninety-six specimens (99%) had an amplification profile compatible with the Omicron variant (Supplemental Table 1). Thirty-two individuals with a positive SARS-CoV-2 RT-qPCR result had a second visit within 7 days, when Ag-RDT from nasal, oral, and saliva specimens were tested again, as well as the RT-qPCR from nasopharyngeal swabs (Figure 3). At the second visit, the positivity rate of RT-qPCR fell to 78.1% (25/32), while the positivity rate of Ag-RDT from nasal specimens fell to 31% (10/32), and to 3% for Ag-RDT from oral or saliva specimens (Figure 3).

**Figure 3.**
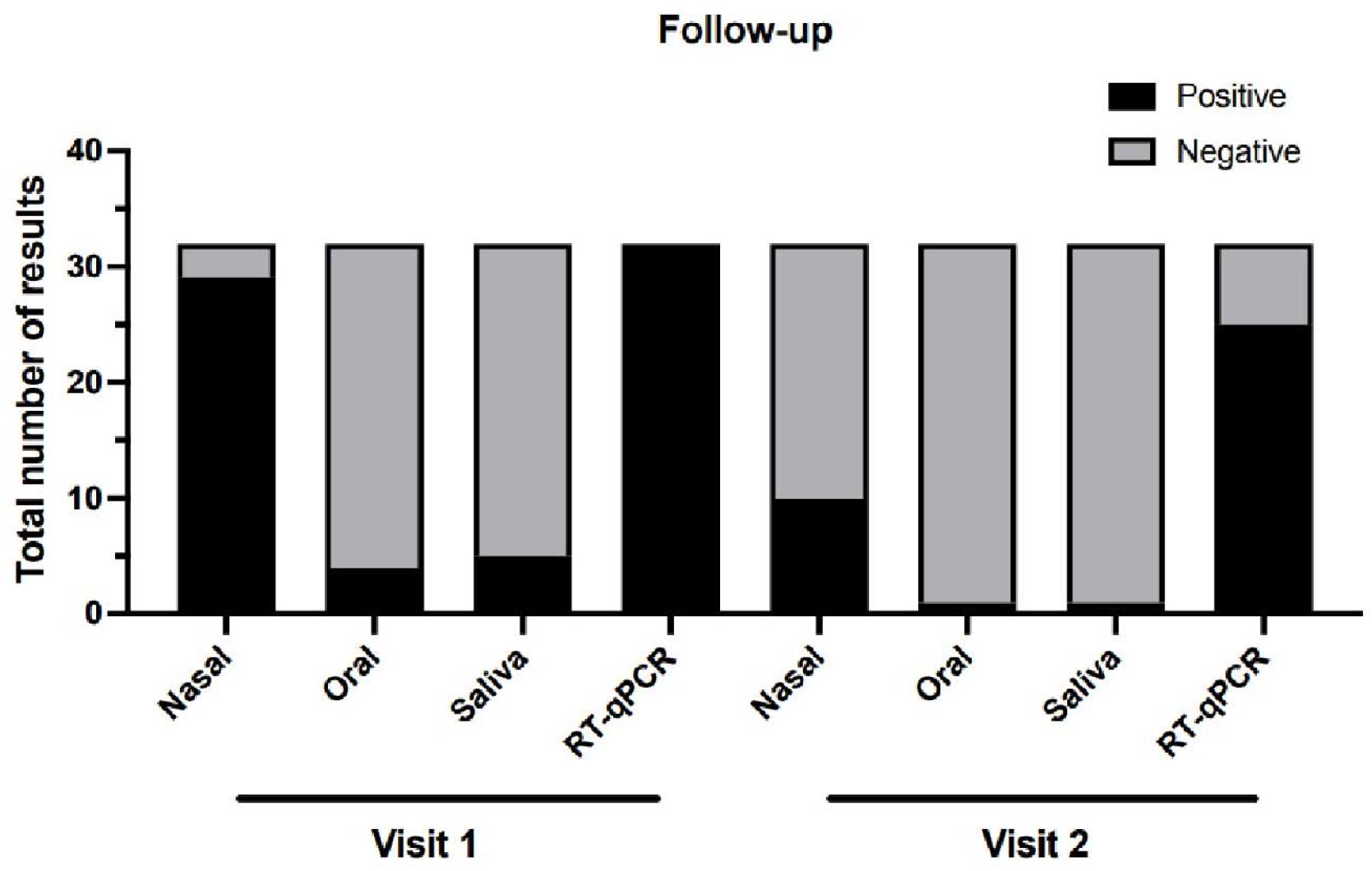
Total number of results obtained by RT-qPCR and Ag-RDT from nasal, oral, and saliva specimens at the first and second visits, in the follow-up group. A total of the 32 patients with a positive SARS-CoV-2 RT-qPCR result were included for follow-up analysis until 7 days after the first test.

Figure 4 shows the concordance among RT-qPCR and Ag-RDTs results during initial and follow-up visits. The positive result observed with Ag-RDT from nasal specimens persisted longer than those detected from oral or saliva specimens. In fact, only nasal Ag-RDT maintained any concordance with RT-qPCR result at the second visit, although significantly lower than that observed at visit 1 (Figure 4).

**Figure 4.**
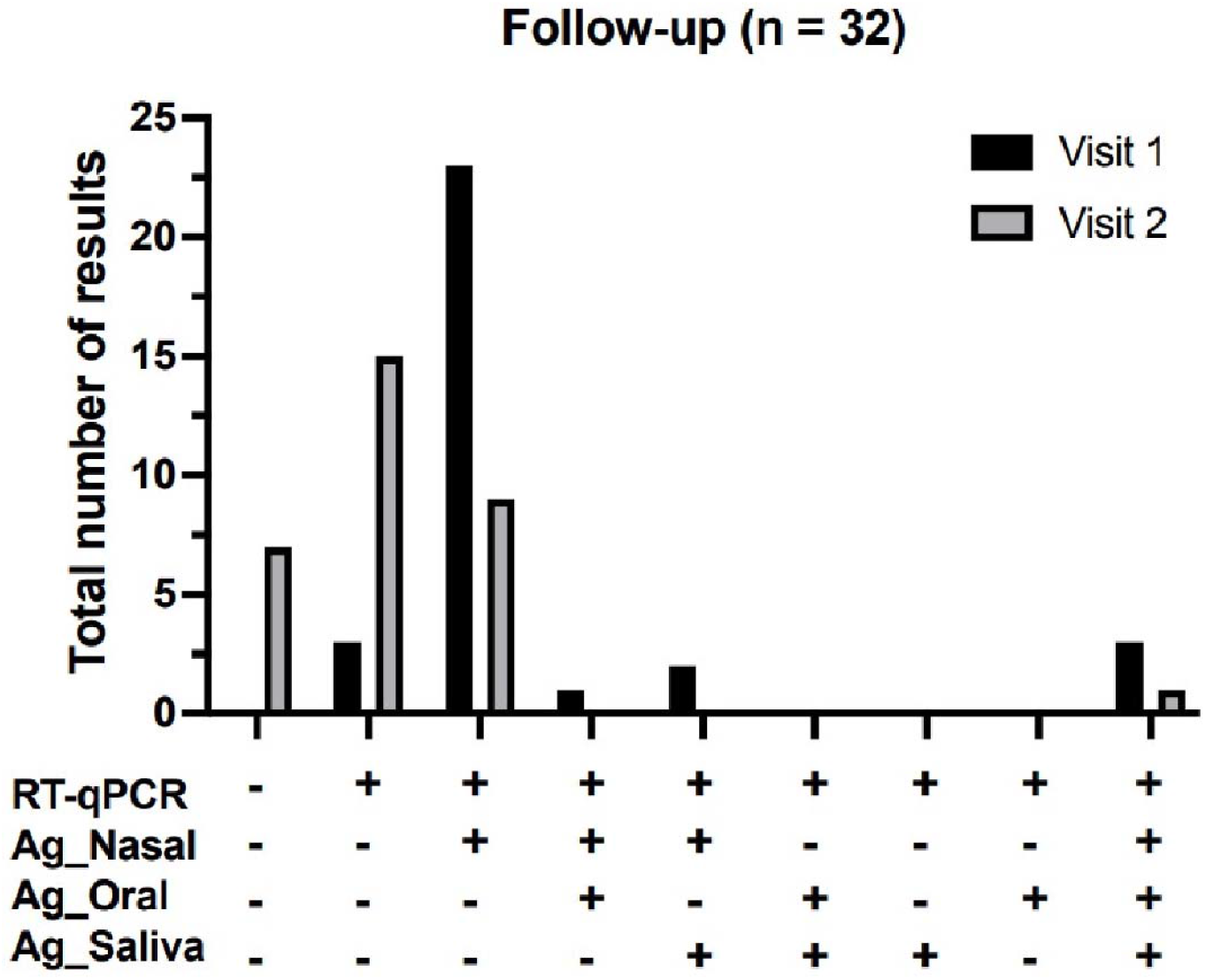
Concordance among SARS-CoV-2 RT-qPCR and Ag-RDT results from nasal (Ag_Nasal), oral (Ag_Oral), and saliva (Ag_Saliva) specimens at the first and second visits in the follow-up group. A total of the 32 patients with a positive SARS-CoV-2 RT-qPCR result were included for follow-up analysis until 7 days after the first test.

## Discussion

Antigen rapid diagnostic tests (Ag-RDT) are an important tool for point of care diagnosis of COVID-19, enabling the implementation of immediate control measures to avoid viral spread. The Panbio™ COVID-19 Ag Rapid Test Device is an Ag-RDT for COVID-19 targeting the viral nucleocapsid protein [18, 19]. It has been previously shown to perform well against SARS-CoV-2 VOCs such as Alfa, Beta, Gamma, and Delta that circulated earlier during the SARS-CoV-2 pandemic [20].

The SARS-CoV-2 variant B.1.1.529 (called Omicron according to WHO nomenclature) originated in South Africa and was first reported on November 24^th^, 2021 [21. The Omicron variant has 50 novel genomic mutations, of which 20 are in the S gene that encodes the Spike protein [22]. Omicron has a higher affinity for human ACE2 receptors compared with the Delta variant, indicating a higher potential for transmission [23,24]. Initial studies showed that infection by the Omicron variant produced less severe disease and significantly reduced the odds of hospital admission compared with earlier variants, such as Delta [25,26]. However, the onset of Omicron circulation raised concerns about Panbio™ COVID-19 Ag RDT performance against this VOC as it incorporated mutations and deletions in the N protein that could potentially affect antigen binding. There is also the possibility of the Omicron VOC modifying its cell tropism and infection kinetics, thus altering the site of viral replication throughout the disease course, thereby changing the appropriate specimen to collect for acute-phase diagnosis [8-12].

A study by Michelena et al., using nasopharyngeal samples (confirmed as Omicron variants by sequencing), showed that the Panbio™ COVID-I9 Ag Rapid Test Device had high sensitivity: 95.6% (Ct ≤20), 92.6% (Ct ≤25), 87.2% (Ct ≤30) and 81.8% (Ct ≤35) compared with nasopharyngeal RT-PCR, with a specificity of 100% [27]. In another study, Deerain et al. evaluated the analytical sensitivity of lateral flow devices against the SARS-CoV-2 Omicron variant using isolates cultured from clinical samples, and demonstrated that the Panbio™ COVID-I9 Ag Rapid Test Device consistently detected a sample (4/4 replicates or 100%) at a concentration of 6.39 log_10_ copies/mL, corresponding to a Ct value of 25.8, underlining the high sensitivity of the test device [28].

A previous report suggested that there are different replication sites for the Omicron VOC in the initial phase of infection, leading to earlier detection in oral or saliva specimens compared with nasal ones [17]. In our study, we examined the sensitivity and specificity of the Panbio™ COVID-19 Ag-RDT in 192 symptomatic individuals up to 7 days after symptom onset, using different specimens including nasal bilateral middle-turbinate swabs, oral swabs, and saliva samples, compared with the gold standard RT-qPCR. The Panbio™ COVID-19 Ag-RDT showed good performance for Omicron detection when using nasal specimens, with 89% sensitivity and 100% specificity. However, oral and saliva specimens performed poorly, with only 12.6% and 22.1% sensitivity, respectively. It is noteworthy that Ag-RDT from oral or saliva specimens alone did not detect any patient with a positive SARS-CoV-2-RT-qPCR result, no matter how early they were tested after symptom onset. RT-qPCR from nasopharyngeal samples and nasal Ag-RDT detected positive cases as soon as 0-1 days after symptom onset.

Our data is in contrast with those reported by Marais 2021 [17], but support previous observations using the Panbio™ COVID-19 Ag RDT for other VOCs detection [29]. On the other hand, Lin 2022 reported no difference between nasal and saliva specimens for Omicron diagnosis [30]. The discrepancy could potentially reflect our patient cohort, where most individuals were fully vaccinated with two doses of COVID-19 vaccine, and a large number of patients having completed the third vaccine dose. This higher immune barrier could modify the natural initial replication site of the Omicron VOC in the oral cavity.

Leftover samples from the Panbio™ COVID-19 Ag RDT were used to perform RT-qPCR using a previously verified procedure [31] The amount of viral RNA obtained by RT-qPCR from the leftover samples, as indicated by Ct values, corroborated the Ag-RDT sensitivity using different specimens. These results showed lower viral concentration in the oral cavity and saliva compartments than in the nasal cavity and nasopharynx, suggesting a virus load 32 times higher in nasal samples than in oral or saliva specimens. Furthermore, the follow-up of a subset of RT-qPCR positive patients showed no variation in the viral load distribution among the upper respiratory tract compartments studied. Additionally, the Ct values obtained in the second test corroborated the better performance of nasal specimens with Ag-RDT than oral or saliva speciemens, supporting the finding that the nasal speciemen is the best specimen type for acute COVID-19 diagnosis using the Panbio™ COVID-19 Ag RDT. These results are consistent with the manufacturer’s instructions that require the use of nasal or nasopharyngeal swab samples with the Panbio™ COVID-19 Ag RDT [18,19]. Studies using alternate specimen types such as oral swabs or saliva samples, which are not recommended in the product *Instructions for Use*, have produced lower sensitivity with the Panbio™ COVID-19 Ag RDT [32,33].

To determine the genotype of SARS-CoV-2 infecting our patient cohort, we used VOC RT-qPCR assay that tests two deletions (Del: S106, G107 and F108; in the ORF1a gene NSP6 and Del: H69 and V70 in the Spike gene) in SARS-CoV-2 genome. The results showed that 96 of the 97 tested specimens (99%) were infected with the Omicron VOC, confirming previous sequence studies in our laboratory showing complete replacement of the Delta VOC by Omicron in the community prior to sample collection for this study.

In conclusion, results from the present study demonstrated that Panbio™ COVID-19 Ag-RDT test performance is unaffected in vaccinated individuals infected with the Omicron VOC. Furthermore, the study demonstrated that nasal specimens are the optimal specimen type for Panbio™ COVID-19 Ag-RDT performance compared with oral and saliva specimens.

## Data Availability

All data produced in the present work are contained in the manuscript.

## Supplementary Information

The VOC RT-qPCR results are presented in Supplemental Table 1.

## Acknowledgements

We would like to thank the entire technical and administrative staff from LVM-IB and CTD-COVID-19/UFRJ for their invaluable support, making this and other studies by our group feasible.

## Funding

This work was sponsored by Abbott Laboratory Grant # CLDG-1001-BR. The funders had no role in the study design, the data collection, the analysis, the interpretation and decision to publish this study.

## Competing interests

The authors declare that there is no conflict of interest to disclose.

## Notes

### Competing Interest Statement

The authors have declared no competing interest.

### Author Declarations

The present study was approved by the local ethics review committee from Clementino Fraga Filho University Hospital (CAAE: 30161620.0.0000.5257) and by the national ethical review board (CAAE: 30127020.0.0000.0068).

