## Supplemental Table 1 for "Evaluation of the Panbio™ COVID-19 Antigen Rapid Diagnostic Test in subjects infected with Omicron using different specimens"

| Sample | RP | SC2-N | Wt Del NSp6 | Wt Del 69,70 | SARS-COV-2 Variant |
|---|---|---|---|---|---|
| 62770 | + | + | NA | NA | Omicron |
| 62771 | + | + | NA | NA | Omicron |
| 62773 | + | + | NA | NA | Omicron |
| 62776 | + | + | NA | NA | Omicron |
| 62782 | + | + | NA | NA | Omicron |
| 62786 | + | + | NA | NA | Omicron |
| 62803 | + | + | NA | NA | Omicron |
| 62814 | + | + | NA | NA | Omicron |
| 62820 | + | + | NA | NA | Omicron |
| 62823 | + | + | NA | NA | Omicron |
| 62827 | + | + | NA | NA | Omicron |
| 62829 | + | + | NA | NA | Omicron |
| 62845 | + | + | NA | NA | Omicron |
| 62849 | + | + | NA | NA | Omicron |
| 62851 | + | + | NA | NA | Omicron |
| 62851 | + | + | NA | NA | Omicron |
| 62853 | + | + | NA | NA | Omicron |
| 62870 | + | + | NA | NA | Omicron |
| 62885 | + | + | NA | NA | Omicron |
| 62886 | + | + | NA | NA | Omicron |
| 62889 | + | + | NA | NA | Omicron |
| 62899 | + | + | NA | NA | Omicron |
| 62915 | + | + | NA | NA | Omicron |
| 62917 | + | + | NA | NA | Omicron |
| 62924 | + | + | NA | NA | Omicron |
| 62932 | + | + | NA | NA | Omicron |
| 62937 | + | + | NA | NA | Omicron |
| 62949 | + | + | NA | NA | Omicron |
| 62956 | + | + | NA | NA | Omicron |
| 62978 | + | + | NA | NA | Omicron |
| 62982 | + | + | NA | NA | Omicron |
| 62993 | + | + | NA | NA | Omicron |
| 63004 | + | + | NA | NA | Omicron |
| 63009 | + | + | NA | NA | Omicron |
| 63010 | + | + | NA | NA | Omicron |
| 63013 | + | + | NA | NA | Omicron |
| 63031 | + | + | NA | NA | Omicron |
| 63035 | + | + | NA | NA | Omicron |
| 63037 | + | + | NA | NA | Omicron |
| 63042 | + | + | NA | NA | Omicron |
| 63045 | + | + | NA | NA | Omicron |
| 63073 | + | + | NA | NA | Omicron |
| 63076 | + | + | NA | NA | Omicron |
| 63077 | + | + | NA | NA | Omicron |
| 63078 | + | + | NA | NA | Omicron |

| | | | | | |
|---|---|---|---|---|---|
| 63088 | + | + | NA | NA | Omicron |
| 63093 | + | + | NA | NA | Omicron |
| 63104 | + | + | NA | NA | Omicron |
| 63105 | + | + | NA | NA | Omicron |
| 63107 | + | + | NA | NA | Omicron |
| 63108 | + | + | NA | NA | Omicron |
| 63111 | + | + | NA | NA | Omicron |
| 63125 | + | + | NA | NA | Omicron |
| 63127 | + | + | NA | NA | Omicron |
| 63128 | + | + | NA | NA | Omicron |
| 63139 | + | + | NA | NA | Omicron |
| 63141 | + | + | NA | NA | Omicron |
| 63144 | + | + | NA | NA | Omicron |
| 63154 | + | + | NA | NA | Omicron |
| 63160 | + | + | NA | NA | Omicron |
| 63165 | + | + | NA | NA | Omicron |
| 63167 | + | + | NA | NA | Omicron |
| 63175 | + | + | NA | NA | Omicron |
| 63179 | + | + | NA | NA | Omicron |
| 63182 | + | + | NA | NA | Omicron |
| 63185 | + | + | NA | NA | Omicron |
| 63190 | + | + | NA | NA | Omicron |
| 63194 | + | + | NA | NA | Omicron |
| 63197 | + | + | NA | NA | Omicron |
| 63198 | + | + | NA | NA | Omicron |
| 63199 | + | + | NA | NA | Omicron |
| 63200 | + | + | NA | NA | Omicron |
| 63203 | + | + | NA | NA | Omicron |
| 63204 | + | + | NA | NA | Omicron |
| 63205 | + | + | NA | NA | Omicron |
| 63207 | + | + | NA | NA | Omicron |
| 63212 | + | + | NA | NA | Omicron |
| 63214 | + | + | NA | NA | Omicron |
| 63218 | + | + | NA | NA | Omicron |
| 63220 | + | + | NA | NA | Omicron |
| 63222 | + | + | NA | NA | Omicron |
| 63225 | + | + | NA | NA | Omicron |
| 63228 | + | + | NA | NA | Omicron |
| 63230 | + | + | NA | NA | Omicron |
| 63231 | + | + | NA | NA | Omicron |
| 63232 | + | + | NA | NA | Omicron |
| 63233 | + | + | NA | NA | Omicron |
| 63234 | + | + | NA | NA | Omicron |
| 63248 | + | + | NA | NA | Omicron |
| 63249 | + | + | NA | NA | Omicron |
| 63259 | + | + | NA | NA | Omicron |

| | | | | | |
|---|---|---|---|---|---|
| 63260 | + | + | NA | NA | Omicron |
| 63261 | + | + | NA | NA | Omicron |
| 63262 | + | + | NA | NA | Omicron |
| 63269 | + | + | NA | NA | Omicron |
| 62795 | + | + | NA | + | Others |
